# Health and Policy Associations of Homelessness in the United States

**DOI:** 10.1101/2022.10.14.22281088

**Authors:** Raghav Awasthi, Vibhor Saxena, Aditya Nagori, Lovedeep Singh Dhingra, Varad Puntambekar, Tavpritesh Sethi

## Abstract

**Importance:** Homelessness is a complex challenge with an estimated yearly economic burden of $6 billion in the United States. Mitigating homelessness requires an understanding of determinants of homelessness, their interaction with health factors, and quantification of impact.

**Objective:** To investigate the health, social and policy factors influencing homelessness in a longitudinal integrative machine learning analysis.

**Data Sources and Study design:** This retrospective longitudinal study integrated Global Burden of Disease (GBD), Health Inequality, and Housing and Urban Development (HUD) datasets for 3131 counties in the United States. We used the disease burden data of 2014, health inequalities data of 2001-2014, and homelessness count of 2015.

**Primary Outcome and Measurement Results:** Homelessness, the burden of disease, health inequalities, economic policies, ethnic, social, and racial factors.

**Methods:** Spearman rank correlation test was performed to check pairwise associations. A unified probabilistic model with temporal causality was fitted using a data-driven structure learning algorithm. The resulting associations adjusted for other variables in the network were quantified using network inference algorithms. Finally, counterfactual analysis was performed to quantify the potential impact of the learned interventions.

**Results:** The total burden of homelessness was significantly (p<0.001) and positively associated with rates of HIV and hepatitis mortality. Inference from the unified probabilistic model indicated that a state with a high hepatitis mortality rate had a 9% higher homelessness. Further, the rate of rheumatic heart disease mortality had a 29% decrease with the provision of shelter in young adults experiencing homelessness (p<0.001). Finally, states with moderate tax progressivity had a mitigating effect on homelessness as compared to both high and low tax progressivity (2% and 5% respectively). We evaluated the counterfactual impact of policy interventions to provide more support to cancer patients to prevent homeless and provision of shelter to prevent rheumatic heart disease mortality in young adults experiencing homelessness.

**Conclusion and Relevance:** Control of infectious diseases and the implementation of tax policies are critical interventions for the reduction of homelessness in the United States.

**Key Points:** Homelessness, Bayesian Network, Counterfactual Analysis

**Question:** What are the health associations and determinants of homelessness in the United States?

**Finding:** In this study on 3131 US Counties, we found infectious diseases mortality and tax progressivity to be strong determinants of homelessness using a Bayesian network model.

**Meaning:** These findings suggest that decreasing the burden of infectious diseases and moderate tax progressivity are vital factors for mitigating homelessness in the United States.

## Introduction

Homelessness is intricately linked to health in the United States. A staggeringly high number of people (567,715) were experiencing homelessness on a single night in 2019, as estimated by the annual point-in-time census of the United States Department of Housing and Urban Development (HUD)^1^. A disproportionate number of mentally or physically challenged individuals are known to be predisposed to homelessness^2,3^ but the association with other health conditions is not completely understood. Up to three million people and about 17 in 10,000 people at any given time are homeless in the United States… Learning to mitigate homelessness is challenging as the associated factors are often complex and self-amplifying, thus making it difficult to distinguish associations from causations. For example, crime is a known effect and a cause of homelessness, with people in jail often finding themselves homeless, and again getting into jail while trying to subsist ^4–6^. Here, we combine different sources of the county and state-level data on homelessness in the United States, the burden of chronic diseases, infections, social and economic disparities, and healthcare indicators. We then learn explainable graphical models with temporal causality and counterfactual analysis to evaluate the impact of the associated factors of homelessness. To the best of our knowledge, this is the first study using explainable machine learning and counterfactual analysis to discover the mitigating factors of homelessness in the United States. Our work combines expert insights from medicine, social health, and demographics to interpret and query the complex model to make relevant inferences.

## Methods

We followed a retrospective longitudinal study design and reported this study according to the EQUATOR’s guidelines on observational studies^7^. The outcome variable was calculated as the state-wise homelessness of the years 2015. County-level Health inequality data from 2001-2014 and County-level burden of disease data of 2014 were taken.

### Study Design

We hypothesized that homelessness is influenced by temporally preceding health-related, social, and policy-related factors. The experiments were set up for this temporally causal question by limiting the homelessness count to the year 2015 alone, but including the disease burden and health inequalities data for years up to 2015. The choice of these years was made due to the availability of this data for these years.

### Data Sources

To learn what would mitigate homelessness, we integrated three distinct county-level and state-level sources of data on (i) state-level homelessness, (ii) county-level mortality due to various diseases, and (iii) county-level health inequity data from The Health Inequality Project.

#### (i) State-level Homelessness

The US Department of Housing and Urban Development (HUD) routinely estimates the number of homeless individuals on any given night. The data used in this study were collected on a night in January 2017 as point-in-time estimates of national and state-level homelessness from 2007-2016. The accompanying Housing Inventory Count (HIC) included data from 2007-2016^8^.

#### (ii) County-level Burden of Disease

These data for the United States were downloaded from the Global Burden of Disease Study (GBD)^9^, a comprehensive regional and global research program of disease burden that assesses mortality and disability from major diseases, injuries, and risk factors. We downloaded county-level 5-yearly mortality risks due to infectious diseases, chronic diseases, respiratory diseases, substance use disorder and intentional injury, cardiovascular disease, and cancer. Although this data included information about life expectancy and age-specific mortality risk from 1980 to 2014, for our analysis, we picked the data from 2014.

#### (iii) County-level Health Inequality

County-level data from The Health Inequality Project (Chetty et al. 2016)^10^ were used in this study. Data contains features representing (1) healthcare, such as quality of preventive care, acute care, percentage of the population insured and Medicare reimbursements, (2) Health behaviors, such as the prevalence of smoking and exercise by income quartiles, (3) Income and affluence of the area, such as median house value and mean household income, (4) Socioeconomic features such as absolute upward mobility, the percentage of children born to single-mothers and crime rate, (5) Education at the K-12 and post-secondary level, school expenditure per student, pupil-teacher ratios, test scores and income adjusted dropout rates, (6) Demographic features, such as population diversity, density, absolute counts, race, ethnicity, migration, urbanization, (7) Inequality indices, such as Gini Index, Poverty rate, Income segregation, (8) Social cohesion indices, such as social capital index, the fraction of religious adherents in the county, (9) Labor market conditions, such as unemployment rate, the percentage change in population since 1980, the percentage change in the labor force since 1980 and fraction of employed persons involved in manufacturing, and (10) Local Taxation.

### Data Integration

We extracted estimates for the age-standardized GBD covariates for both sexes for the year 2014. These county-level mortality indicators were merged with the county-level health inequality data from 2001 to 2014. Homelessness data were available as a part of continuum-of-care (CoC) programs, each of which spans multiple counties within a state in the US. We summarised all the homelessness-related variables at the CoC level & state level and filtered for the year 2015. These were then merged with county-level GBD and health inequality data until the year 2014, as described above.

### Imputation and Discretisation of Data

Some data were missing in the health inequality variables. We removed the variables in which more than 10% of data was missing and imputed missing values of the data using state of the art imputation technique, missforest^11^. We then performed the discretization of continuous variables into three intervals (labeled as low, medium, and high). This was done using an in-house code pipeline written in R that runs k-means, frequency-based, quantile and uniform-interval based methods (in that order of preference) for each variable. The number of discrete classes was fixed at three for ease of interpretation and decisions.

### Statistical Analysis

Statistical analyses to explore associations and potential causation were carried out using R language and environment for statistical computing^16^. A pairwise Spearman Correlation test was first performed to explore the relationship of homelessness variables with disease mortality and socioeconomic variables.

Next, a data-driven structure learning analysis was carried out using Bayesian Networks in order to learn a joint probabilistic model instead of pairwise correlations., A robust consensus network was inferred by learning a sufficiently high number odd number(501) of Bayesian networks which were bootstrapped from data and ensemble-averaged. The networks were learned using Hill climbing Search Algorithm^12^ and BIC (Bayesian Information Criterion) Score Function^13^. Bayesian networks are directed acyclic graphs and since variables from the future cannot influence the variables in the past, blacklisting of variables has been done while learning networks that are reversed in a temporal proceeding (, that is, the links from 2015 to 2014). The parents of homelessness nodes were specified as potentially mitigating influences, Third, from the Bayesian network, we determined the Markov blanket^14^ of the homelessness variables, which are more important for mitigating homelessness. Then we analytically calculated the conditional probability of determined Markov blanket variables and inferred the pattern for change in homelessness when we change Markov blanket variables moderate to low, moderate to high, and low to high.

Next, we imagined the hypothetical situation of the variables (Derived from Markov Blanket of Bayesian Network) and performed **counterfactual analysis**^15^ to answer the hypothetical ‘what if’ questions. For example, what would be the homelessness of a high tax state if they would have prevailed the characteristics of a moderate tax state (Counterfactual count). We calculated the difference between the original count of homelessness and the counterfactual count of homelessness in ten disjoint quantiles called quantile effects. A positive quantile effect reflects a decrease in homelessness upon supplementing with a changing tax structure and a negative quantile effect reflects the increase in homelessness upon providing with changing the tax structure.

## Results

### From Associations to Directed Relations to Counterfactuals

#### 1 Associations between Disease Burden, Homelessness, and Taxation in the United States

In the Spearman Correlation test, we observed that the counties with a diverse population (Hispanic, Foreign-born, and Black), numerous uninsured people, and mortality rate of infectious diseases like Hepatitis, Tuberculosis, HIV, and Acute Lymphoid Leukemia surging in the area, are positively correlated with Homelessness, and all variables exhibit positive correlation (correlation>=0.40, p-value <.001) with Unsheltered Homelessness which is 22.3% of the total homeless in all states. However, the mortality rates of Rheumatic heart disease, Pneumoconiosis, Chronic Respiratory diseases are found to be negatively correlated (correlation<-0.40, p-value <.001) with sheltered Parenting Youth Homelessness. The tax progressivity of the states showed a nonlinear effect of correlation among homelessness variables such that is positively correlated with sheltered homelessness variables and negatively with unsheltered homelessness variables.[Figure 1]

**Figure 1:**
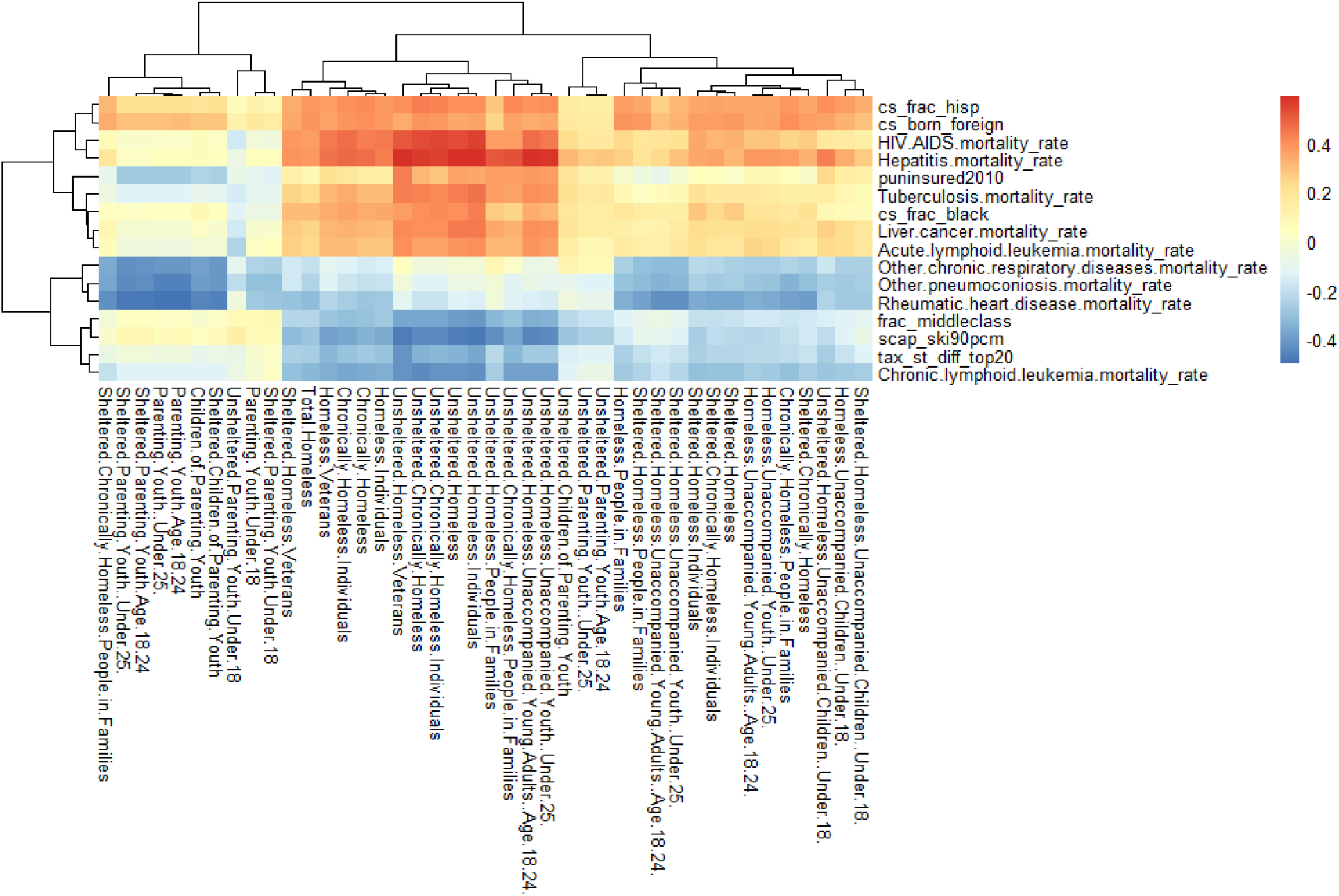
Heat map of Spearman Correlation between Global Burden of Disease (2014), Health Inequality (2001-2014), and Homelessness (2015). Variables of GBD and HI are shown, which have a magnitude of correlation of more than 0.4.

#### 2 Directed Relationships and Statistical Inference: Bayesian Network Analysis

From 501 Bootstrapped ensemble-averaged Bayesian network[Figure 2] by choosing the majority of voting, tax progressivity comes out as the strongest influencer Homeless along with multiple Disease Mortality in Bayesian Network. Hepatitis Mortality is a parent node of unsheltered children of parenting youth [Table 1], Tax progressivity is the parent node of “Unsheltered homelessness”, “unsheltered homelessness in young adults”, and sheltered chronically homeless people., Tax Progressivity, Rheumatic Heart disease, Diarrhea, Hepatitis, larynx cancer, kidney cancer was found in the Markov blanket variables of homelessness variables, in the Bayesian network model we conditioned that edge strength and direction strength must be greater than 0.51 and then calculated the differences in conditional probability on homelessness variables when tax progressivity is high [2.41,7.22] and moderate [0.639,2.41] similarly tax structure is regressive [0,.639] to tax structure is moderate, same the difference in probability of homelessness variables rate of Disease mortality is high and Rate of disease Mortality is low.[Figure 3]

**Table 1.** Parent-Child nodes of the Bayesian Network with edge strength greater than 60%.

| Parent | Child [Edge Strength] |
| --- | --- |
| Tax Progressivity | • Unsheltered Homelessness [100%]<br>• Unsheltered Children of Parenting Youth [64%]<br>• Sheltered Homelessness Unaccompanied Young Adults Age 18-24 [60.04%]<br>• Sheltered Chronically Homeless People in the Family [100%]<br>• Homeless Unaccompanied Young Adults Age 18-24 [98 %] |
| Rheumatic Heart Disease Mortality Rate | Sheltered Homeless Unaccompanied Young Adults Age 18-24 [64%] |
| Diarrhea Mortality Rate | Sheltered Chronically Homeless [90%] |
| Hepatitis Mortality Rate | Unsheltered Children of Parenting Youth [74%] |
| Kidney Cancer Mortality Rate | Unsheltered Children of Parenting Youth [72%] |
| Larynx Cancer Mortality Rate | Sheltered Homelessness [81%] |

**Figure 2:**
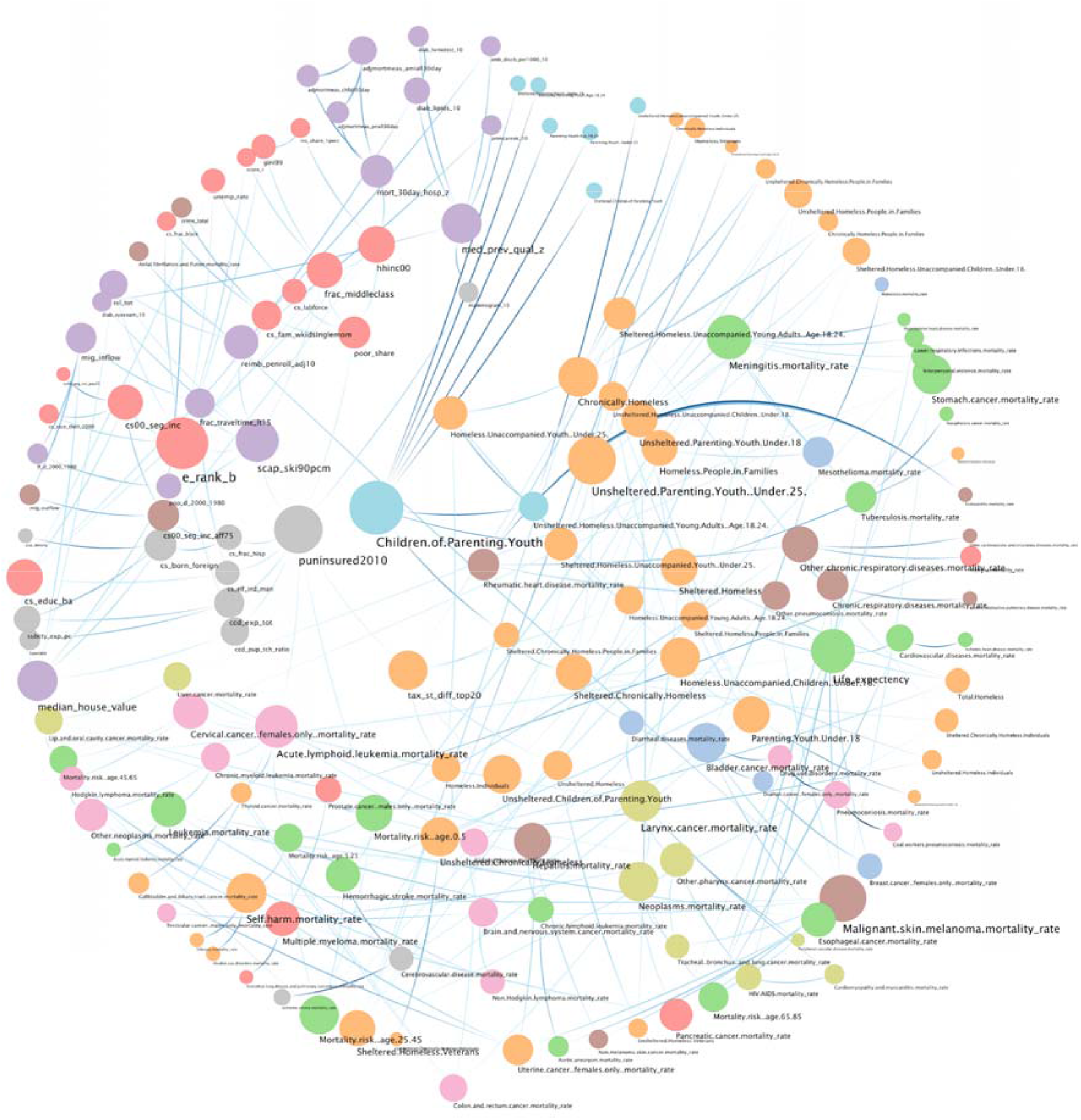
Ensemble network learned upon Global Burden of Disease (GBD) Data, Health Inequalities (HI) Data, and Homelessness Data-The majority-voted structure from 501 bootstrap structure-learning iterations used hill-climbing search along with Bayesian Information Criterion as the scoring function.

**Figure 3:**
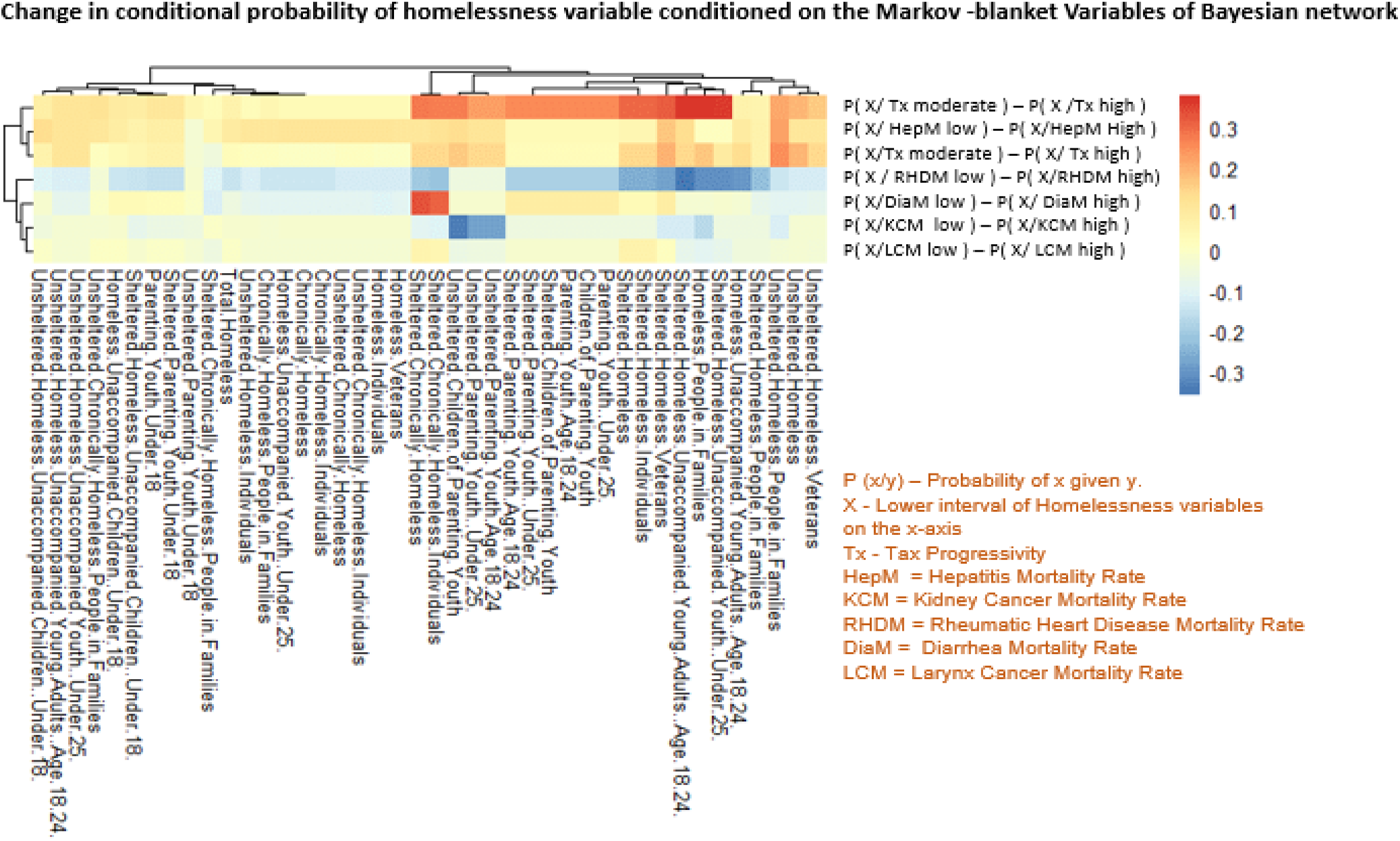
Heat Map of Difference in Conditional Probability of the Variable given on the x-axis with respect to the Change in the Condition given on the y-axis.

#### 3 Counterfactual Analysis

We observed a positive quantile effect at the median quantile in hepatitis mortality rate and tax progressiveness. We found a difference of 6394 homeless people with 95% CI [5941.71,6846.92] if the states with high hepatitis mortality rate prevailed the characteristics of a state with low hepatitis mortality rate. Similarly, we observed a difference of 1989 homeless people with 95%CI [-122.46, 4099.99] if the states with high progressive tax would have prevailed the characteristics of a state that is the moderately tax-progressive state. A negative quantile effect is observed in Rheumatic heart disease mortality rate, kidney cancer mortality rate, and diarrhea mortality rate, as shown in. [Table 2.]

**Table 2:** Counterfactual Effect of Health and Policy indicators on Homelessness.

| Indicator | Counterfactual condition | Quantile effect [95 % Confidence Interval] |
| --- | --- | --- |
| Hepatitis Mortality | High to Low | 6394.32 [5941.71,6846.92] |
| Tax Progressivity | Progressive to Moderate | 1988.76l [ -122.46, 4099.99] |
| Tax Progressivity | Regressive to Moderate | 850.22[-1009,2709] |
| Rheumatic Heart Disease Mortality | High to Low | -3878.91[-4263.05,-2221.82] |
| Kidney Cancer Mortality | High to Low | -3655.13[ -4782.68,-2527.57] |
| Diarrhea Mortality | High to Low | -2030.19[-3001.62,-1058.76] |

## Discussion

Homeless people are the most vulnerable due to, unavailability of accessible and affordable healthcare. Homeless people are at the highest risk of the spread of infectious diseases, premature mortality, mental disorders, and unintentional injuries among many others.^17^, To the best of our knowledge, this is the first application of graphical models in large multivariate datasets to learn factors that were responsible for the mitigation of homelessness in the United States. The probabilistic joint distribution revealed tens of meaningful insights about disparities in insurance, cancer, diversity, and race that lead to homelessness in the United States. Our model has also captured key influencers such as the tax progressiveness of states and the potential need to adopt a moderately progressive tax structure. Our robust multi-step analysis is also able to capture all linear and nonlinear trends of association. Results were consistent from correlation values to directed relation to counterfactual effect. Correlation values revealed that there is a linear association of homelessness with mortality and socioeconomic factors, highly correlated variables like hepatitis mortality rate, diarrhea mortality rate, Rheumatic heart disease mortality, tax progressivity comes out to be the most influential variable in the Bayesian network; and counterfactual analysis verified the claim of Bayesian Network Inferences. Hepatitis mortality was found positively correlated with homelessness which shows that control in hepatitis mortality can mitigate homelessness but also some cancer mortality and rheumatic heart disease mortality are negatively correlated with homelessness which shows that homeless people are at high risk of disease if they die due to these diseases then mortality rate increases and homelessness decreases. The key strength of our paper is the robust and counterfactual analysis to identify the mitigators of homelessness. We also published a dashboard of our network analysis, which is useful for enabling decisions.

Our study has several limitations: (i) unavailability of the latest data, and (ii) extensive inclusion of environmental and educational indicators. Even though the health inequality data have an exhaustive set of socioeconomic and demographic variables, the inclusion of more variables would lead to a more comprehensive analysis.

## Data Availability

Publicly Available

https://www.hud.gov/press/press_releases_media_advisories/hud_no_21_041

https://www.healthdata.org/gbd/data-visualizations

